# Association of Mobile Stroke Unit Care and Spending, Utilization, and Mortality in New York City

**DOI:** 10.1101/2024.02.13.24302801

**Authors:** Dhruv Khullar, Babak B. Navi, Saad Mir, Matthew E. Fink, Luisa Tinapay, Glenn Asaeda, Rainu Kaushal, Eshani Pareek, Amelia M. Bond

**Affiliations:** Department of Population Health Sciences, Weill Cornell Medicine, New York, NY; Department of Medicine, Weill Cornell Medicine, New York, NY; Department of Neurology and Feil Family Brain and Mind Research Institute, Weill Cornell Medicine, New York, NY; Department of Emergency Medicine, Weill Cornell Medicine, New York, NY; Fire Department of New York, Brooklyn, NY

## Abstract

**Background:** Transport by mobile stroke units (MSUs), which provide access to computed tomography scanning and intravenous blood pressure medications and thrombolytics, reduces time to treatment and may improve short-term functional outcomes for patients with acute stroke. The longer-term clinical and financial impacts remain incompletely understood.

**Objective:** Determine whether MSU care is associated with better health, utilization, and spending outcomes for patients with suspected acute stroke.

**Design:** Retrospective, observational study of Medicare patients transported by MSUs versus traditional ambulances in New York City, from October 2016 to December 2019.

**Eligibility:** 167 Medicare patients with suspected acute stroke transported by MSU and 2,518 propensity-score matched controls.

**Main Outcomes:** Primary outcomes included length of stay and discharge destination at index hospitalization, as well as risk of repeat hospitalization, number of emergency department visits, total costs of care, and mortality at 1 year.

**Results:** Of 167 patients (mean age, 79.9 years;56.3% women) transported by a MSU for suspected acute stroke, 61.1% had an ischemic stroke/TIA, 7.8% had an intracerebral hemorrhage, and 31.1% had a stroke mimic or other diagnosis. Compared to propensity-score matched control patients, MSU patients experienced similar lengths of stay (5.9 vs 6.7 days,p=0.13) and were similarly likely to be discharged to a skilled nursing facility (15.6% vs 15.1%,p=0.86). They had clinically but marginally significant lower rates of mortality at 1 year (21.6% vs 28.4%; difference, 6.8 percentage points [95% CI -13.3 to 0.3,p=0.058). They had similar rates of any repeat hospitalization (24% vs 23.2%,p=0.82) and ED visits without hospitalization (14% vs 12%,p=0.86), and there were no significant differences in total spending or specific types of spending.

**Conclusions:** In this study of patients presenting with suspected acute stroke in New York City, transport by MSUs, compared with traditional ambulances, was associated with a trend toward lower mortality at 1 year. Prospective trials and replication in other regions are warranted.

## Introduction

Rapid initiation of treatment is critical for good clinical outcomes for patients with acute stroke.^1,2^ Many patients do not receive prompt treatment, however, often because of delays in reaching the hospital.^3^ Mobile stroke units (MSUs) are ambulances equipped with clinical staff and computed tomography scanners that expedite treatment for acute stroke compared to transport via traditional emergency medical services (EMS). Research suggests that MSU care is associated with faster initiation of intravenous tissue plasminogen activator (t-PA) thrombolysis, including within the so-called Golden Hour (i.e., within 60 minutes of the onset of symptoms), and better functional outcomes, as measured by the modified Rankin scale (mRs) at 90 days, for patients with ischemic stroke.^4–9^

The impact of MSU care on other health and spending outcomes, such as hospital length of stay, acute care utilization, total costs of care, and mortality, is less well studied. Moreover, many studies assess outcomes during the index hospitalization and for 3 months after, but do not analyze long-term effects of MSU care. Finally, more research is needed to understand the impact of MSU care on different patient populations, given that stroke outcomes are known to vary substantially by age and other patient characteristics, and across different localities, because the receipt of timely stroke care can vary by traffic patterns, public awareness, and health system resources.^10–12^

In this study, we examined outcomes for Medicare patients presenting with symptoms of acute stroke who were transported by MSUs versus those using traditional transport in New York City. Specifically, we sought to determine the association of MSU care with short-term hospital outcomes, such as length of stay and discharge destination, as well as longer-term health and spending outcomes, including health care utilization, spending, and mortality at 1 year.

## Methods

This study was approved by the Weill Cornell Institutional Review Board and informed consent was waived because only historical data with minimal risk of harm were used. This study followed the Strengthening the Reporting of Observational Studies in Epidemiology (STROBE) reporting guideline.^13^

### Data and Sample

The main data sources included the New York City MSU registry, 100% 2016-2020 Medicare fee-for-service claims, 2019 5-year American Community Survey, as well as electronic medical records from two New York Presbyterian Manhattan hospitals through the Tripartite Request Assessment Committee (TRAC) network.^14^ Using Medicare claims, we identified all admissions between October 2016 and December 2019 for patients aged 65 years or older occurring in a hospital serviced by a New York MSU with an admitting or primary discharge diagnosis of acute stroke or a relevant stroke mimic. To identify our case population, we next used the MSU registry to identify all patients transported via the New York MSU, which serves the boroughs of Manhattan, Queens, and Brooklyn, for suspected acute stroke between October 2016 and December 2019. We then linked the MSU transports of patients 65 years and older to the set of acute stroke or stroke mimic inpatient Medicare claims using age, sex, date of transport, and admitting hospital with a multi-step approach (eMethods in **Supplement**). Further, we used the American Community Survey to link patients’ zip-code level demographic information to Medicare claims. Finally, we used TRAC data, that contained data for a secondary clinical outcome, to link patients with a hospitalization at two New York Presbyterian Manhattan hospitals between October 2016 and December 2019 to hospitalizations in Medicare claims using unique patient identifiers including Social Security Number, Health Insurance Claim Number, or Medicare Beneficiary Identifier.

Admissions with a length-of-stay less than 1 day were excluded. For patients with multiple admissions during the time frame, only the index admission was included.

### Outcomes

Our primary outcomes were short-term hospital outcomes as well as longer-term health, utilization and spending outcomes. Short term hospital outcomes included the length-of-stay of the index hospitalization and whether a patient was discharged home. Longer term utilization measures included whether a patient had any inpatient or emergency department visits within one year of discharge. We also examined total spending within one year of discharge as well as total spending by broad categories including inpatient, outpatient, post-acute care and hospice. Finally, we measured one long-term health outcome – mortality within one year post discharge.

The secondary outcome available for a subset of patients admitted to two Manhattan hospitals was a modified Rankin Score (mRS) in electronic medical records collected at the time of discharge from an index admission.

### Other Covariates

Patient demographic information included age, race, and sex. Two residential demographic variables were added from the American Community Survey based on a patient’s zip code: proportion with high school diploma and median household income.

Clinical characteristics prior to hospitalization included residence at a care facility, Hierarchical Condition Category (HCC) risk score as well as indicators for whether a patient had atrial fibrillation, diabetes, or hypertension. Residence at a care facility immediately prior to an index admission was constructed using MDS Nursing Home Assessment data and identified based on an assessment within 90 days prior to stroke hospitalization with no discharge to another facility. The HCC risk score was constructed using Medicare claims data the year prior to a patient’s index admission. Indicators for chronic conditions were obtained through the Chronic Conditions segment of the Master Beneficiary Summary File the year prior to a patient’s index admission. Based on the admitting or primary discharge diagnosis from the index hospitalization claim, hospitalizations were categorized into stroke types as ischemic stroke/TIA, intracerebral hemorrhage (ICH), subarachnoid hemorrhage (SAH), or other diagnosis, which included stroke mimics. For the MSU group, the working diagnosis of the treating attending neurologist at the time of MSU transport is also provided.

For a sensitivity analysis, we constructed three measures of a patient’s medication use up to one year prior to admission using Medicare Part D claims. Measures included any use of statins, anticoagulants, and insulin. For a secondary outcome (mRS) analysis, patients’ NIH Stroke Scale (NIHSS) was also collected from electronic medical records at the time of admission during the index hospitalization.

Data on intravenous tPA use was collected. For patients transported via an MSU ambulance, tPA administration was documented in the registry. For patients not transported via an MSU ambulance, tPA administration was determined by ED claims data in the MedPAR or outpatient file, using codes identified in previous literature.^15,16^

### Analysis

MSU and control admissions were propensity score matched to account for potentially confounding variables. The propensity score was constructed using logit regressions where MSU treatment was the dependent variable. Propensity score matched controls were identified on a 1:20 basis using Mahalanobis distance matching. The 1:20 match rate was based on a prespecified power calculation for a greater than 10% change in discharge to home following admission. Controls were matched by patient age, sex, Hierarchical Condition Category (HCC) risk score, chronic condition indicators, two residential demographic variables, as well as year and quarter of admission. Due to the sample size of patients with mRS, the match rate was 1:5 for this secondary outcome. All controls in the main analysis were used in the secondary analysis with the additional NIHSS adjustment. Admissions were separately matched for admitting hospital, stroke type, and whether a patient was transported from a care facility and then combined for analysis. We tested the balance of the sample by comparing covariates across treatment and control admissions using t-tests or tests of proportion. We also compared outcomes across treatment and control admissions using t-tests or tests of proportion. Administration of tPA was not used for matching because it is thought to be a key mechanism through which outcomes may improve for MSU vs non-MSU patients.

As tests of sensitivity, we performed three more analyses. First, we limited the set of MSU treatment admissions to those that satisfied a stricter set of match criteria between the MSU registry and Medicare claims (eMethods in **Supplement**). Second, focusing only on patients with Part D, we propensity score matched using three additional controls that measured a patient’s medication use prior to admission. Third, we added race as a covariate in the propensity for matching.

All statistical analyses were performed using Stata, version 17.0 (StataCorp), and 2-sided P<0.05 was considered statistically significant. The study was deemed exempt from review by the institutional review board at the Biomedical Research Alliance of New York.

## Results

Between October 2016 and December 2019, 294 patients aged 65 years and older were transported by a New York City MSU, including 167 Medicare patients who met the study’s inclusion criteria (**Figure 1**). The mean age was 79.9 years; 56.3% of patients were women and 61.1% had a stroke type of ischemic stroke/TIA. MSU patients and propensity-score matched control patients (N=2518) were similar on a broad range of clinical and demographic characteristics, including age, sex, comorbidities, and type of stroke (**Table 1**). MSU patients were significantly more likely to receive tPA relative to matched patients (49.9% vs 9.4%, p<0.001). For the MSU patients, the working diagnosis of the attending neurologist at the time of MSU transport were ischemic stroke (n=130, 77.8%), acute encephalopathy (n=17, 10.1%), ICH (n=15, 9.0%), TIA (n=2, 0.6%), SAH (n=1, 0.3%), seizure (n=1, 0.03%), and recrudescence of old stroke (n=1, 0.03%).

**Table 1:**
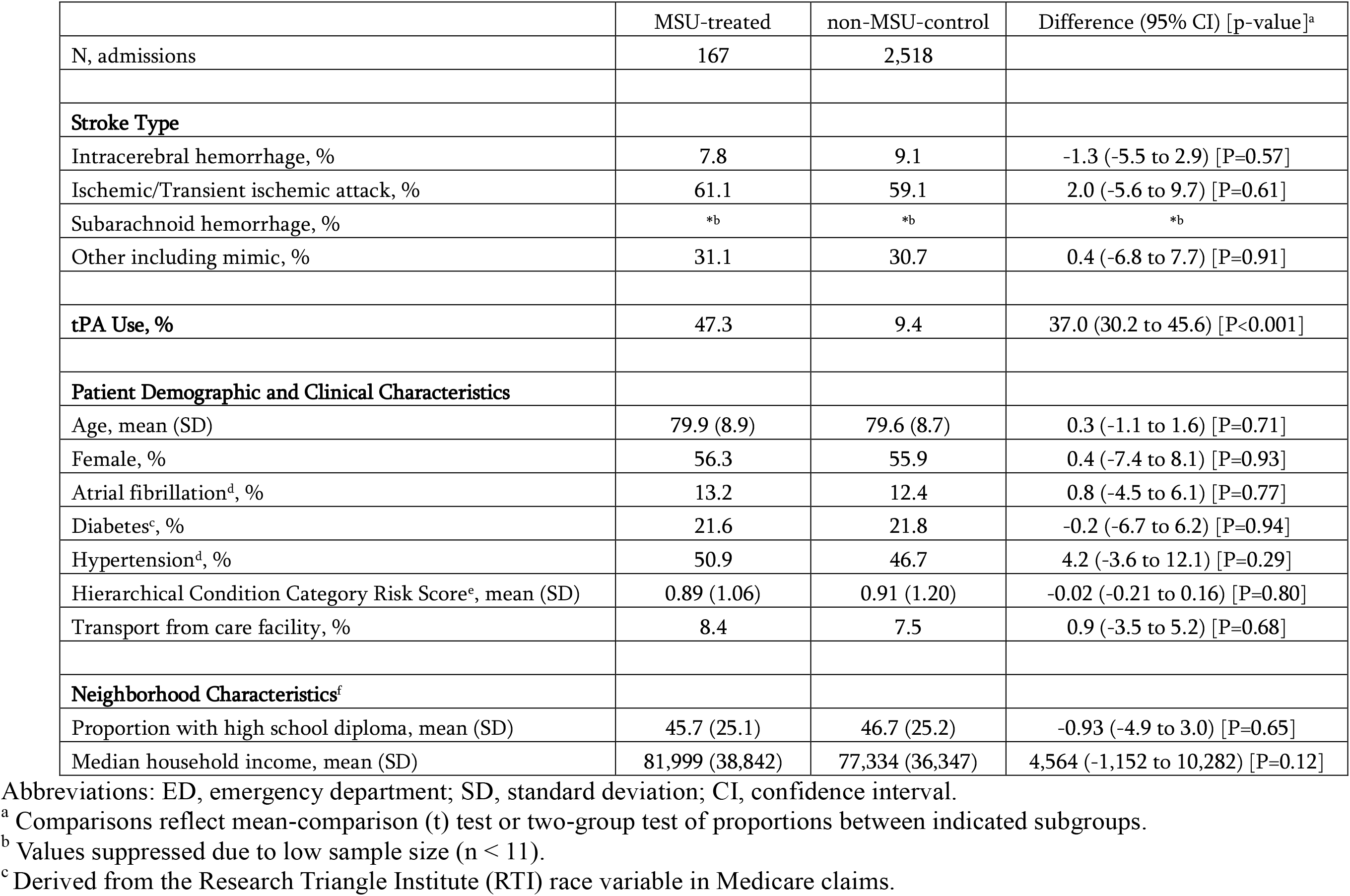
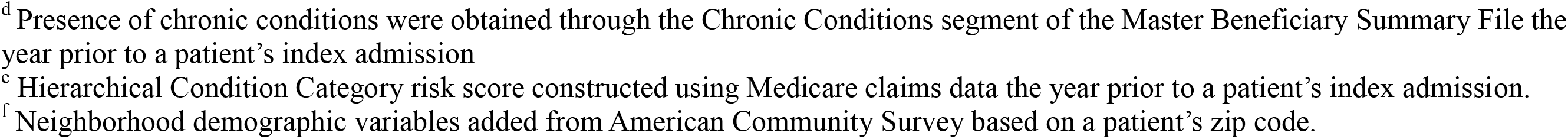
Demographic and Clinical Characteristics by MSU Treatment.

|  | MSU-treated | non-MSU-control | Difference (95% CI) [p-value]<sup>a</sup> |
| --- | --- | --- | --- |
| N, admissions | 167 | 2,518 |  |
| **Stroke Type** |  |  |  |
| Intracerebral hemorrhage, % | 7.8 | 9.1 | -1.3 (-5.5 to 2.9) [P=0.57] |
| Ischemic/Transient ischemic attack, % | 61.1 | 59.1 | 2.0 (-5.6 to 9.7) [P=0.61] |
| Subarachnoid hemorrhage, % | *b | *b | *b |
| Other including mimic, % | 31.1 | 30.7 | 0.4 (-6.8 to 7.7) [P=0.91] |
| **tPA Use, %** | 47.3 | 9.4 | 37.0 (30.2 to 45.6) [P<0.001] |
| **Patient Demographic and Clinical Characteristics** |  |  |  |
| Age, mean (SD) | 79.9 (8.9) | 79.6 (8.7) | 0.3 (-1.1 to 1.6) [P=0.71] |
| Female, % | 56.3 | 55.9 | 0.4 (-7.4 to 8.1) [P=0.93] |
| Atrial fibrillation<sup>d</sup>, % | 13.2 | 12.4 | 0.8 (-4.5 to 6.1) [P=0.77] |
| Diabetes<sup>c</sup>, % | 21.6 | 21.8 | -0.2 (-6.7 to 6.2) [P=0.94] |
| Hypertension<sup>d</sup>, % | 50.9 | 46.7 | 4.2 (-3.6 to 12.1) [P=0.29] |
| Hierarchical Condition Category Risk Score<sup>e</sup>, mean (SD) | 0.89 (1.06) | 0.91 (1.20) | -0.02 (-0.21 to 0.16) [P=0.80] |
| Transport from care facility, % | 8.4 | 7.5 | 0.9 (-3.5 to 5.2) [P=0.68] |
| **Neighborhood Characteristics<sup>f</sup>** |  |  |  |
| Proportion with high school diploma, mean (SD) | 45.7 (25.1) | 46.7 (25.2) | -0.93 (-4.9 to 3.0) [P=0.65] |
| Median household income, mean (SD) | 81,999 (38,842) | 77,334 (36,347) | 4,564 (-1,152 to 10,282) [P=0.12] |

Abbreviations: ED, emergency department; SD, standard deviation; CI, confidence interval.
<sup>a</sup> Comparisons reflect mean-comparison (t) test or two-group test of proportions between indicated subgroups.
<sup>b</sup> Values suppressed due to low sample size (n < 11).
<sup>c</sup> Derived from the Research Triangle Institute (RTI) race variable in Medicare claims.
<sup>d</sup> Presence of chronic conditions were obtained through the Chronic Conditions segment of the Master Beneficiary Summary File the year prior to a patient's index admission
<sup>e</sup> Hierarchical Condition Category risk score constructed using Medicare claims data the year prior to a patient's index admission.
<sup>f</sup> Neighborhood demographic variables added from American Community Survey based on a patient's zip code.

**Figure 1:**
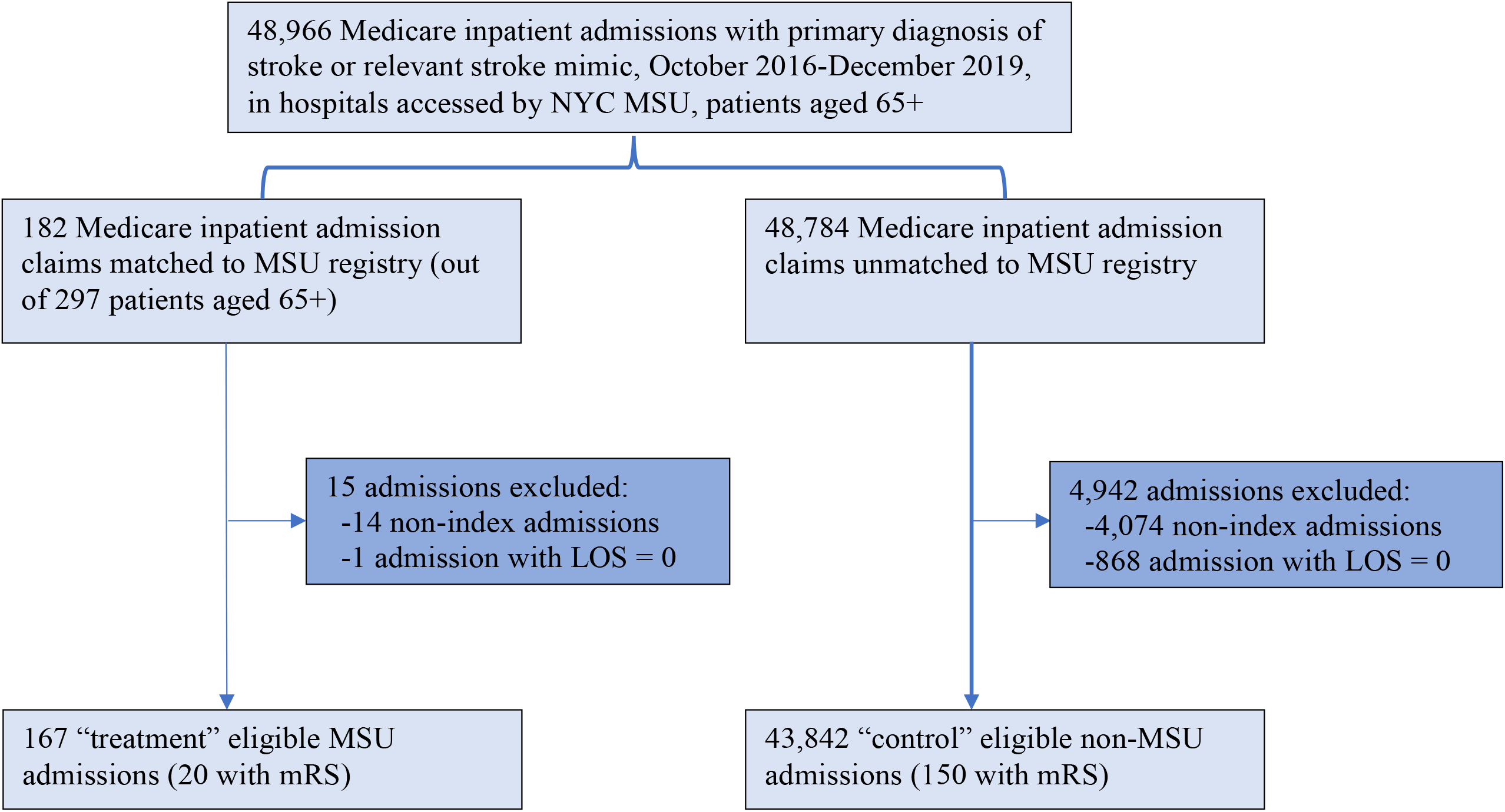
Sample Flow Chart.

**Table 2** presents outcomes during index hospitalization. Compared to patients transported by traditional ambulances, MSU patients had similar lengths of stay (5.9 vs 6.7 days, p=0.13), and were discharged to skilled nursing facilities at similar rates (15.6% vs 15.1%, p=0.86).

**Table 2:** Clinical, Utilization, and Spending Outcomes by MSU Treatment.

|  | MSU-treated | non-MSU-control | Difference (95% CI) [p-value]<sup>a</sup> |
| --- | --- | --- | --- |
| N, admissions | 167 | 2,518 |  |
| **Clinical** |  |  |  |
| Length of stay, mean (SD) | 5.86 (5.54) | 6.68 (6.71) | -0.81 (-1.85 to 0.23) [P=0.13] |
| 1 year Mortality, % | 21.6 | 28.4 | -6.8 (-13.3 to 0.3) [P=0.06] |
| Discharge to SNF, % | 15.6 | 15.1 | 0.5 (-5.2 to 6.2) [P=0.86] |
| **Utilization, 1 year post discharge** |  |  |  |
| Any inpatient admission, % | 24.0 | 23.2 | 0.8 (-5.9 to 7.4) [P=0.82] |
| Any emergency department visit not resulting in inpatient admission, % | 13.8 | 11.6 | 0.5 (-5.2 to 6.2) [P=0.86] |
| **Spending, 1 year post discharge<sup>b</sup>** |  |  |  |
| Total, mean (SD) | 33,526 (48,369) | 32,191 (50,317) | 1,335 (-6,527 to 9,197) [P=0.74] |
| Inpatient, mean (SD) | 14,463 (31,171) | 14,815 (31,584) | -352 (-5,123 to 4,419) [P=0.95] |
| Outpatient, mean (SD) | 3,702 (8,902) | 2,944 (7,459) | 758 (-426 to 1,941) [P=0.21] |
| Post-acute, mean (SD) | 12,578 (25,264) | 13,371 (27,104) | -794 (-5,021 to 3,434) [P=0.71] |
| Hospice, mean (SD) | 1,274 (4,619) | 1,157 (4,204) | 117 (-545 to 780) [P=0.73] |

Abbreviations: ED, emergency department; SD, standard deviation; CI, confidence interval.
<sup>a</sup> Comparisons reflect mean-comparison (t) test or two-group test of proportions between indicated subgroups.
<sup>b</sup> Spending does not include cost of index admission.

**Table 2** also presents health, utilization, and spending outcomes at 1 year after hospital discharge. Compared to patients transported by traditional ambulances, MSU patients had clinically lower rates of mortality, although the difference was marginally statistically significant (21.6% vs 28.4%, p=0.058). They were similarly likely to experience a repeat hospitalization within 1 year of discharge (24.0% vs 23.2%, p=0.82), and had similar likelihood of ED visits without inpatient admission (13.8% vs 11.6%, p=0.86). Overall spending at 1-year post-discharge was also similar between MSU and control patients ($33,526 vs $32,191, p=0.74), and there were no significant differences by type of spending (inpatient, outpatient, post-acute, or hospice).

Results were broadly similar in sensitivity analyses. However, in analysis with a more conservative set of treatment observations (eMethods and eTable 1 in **Supplement**), MSU patients had significantly lower lengths of stay during the index hospitalization and the mortality difference, while large, was again only marginally significant (eTable 2 in **Supplement**). In an analysis of patients with Medicare Part D and controlling for prior medication use, MSU patients had significantly more outpatient spending and the mortality difference was also large, but not statistically significant (eTable 3 and 4 in **Supplement**). Finally, in analysis using race as an additional demographic covariate, the mortality difference between MSU and control patents was large, but marginally statistically significant (eTable 5 and 6 in **Supplement**).

In a secondary analysis examining patients for whom mRS were available at time of discharge, we found that MSU patients had similar mRS relative to non-MSU patients (2.20 vs 2.48, p=0.56) (eTable 7 and 8 in **Supplement**).

## Discussion

In this retrospective, observational study of patients presenting with suspected acute stroke who were transported by MSUs vs traditional transport in New York City, MSU care was associated with a trend toward lower mortality at 1 year, although this finding did not reach statistical significance, possibly owing to the sample size. MSU patients experienced shorter lengths of stay during the index hospitalization, although this finding did not reach statistical significance. Other outcomes, including utilization of inpatient and ED care, as well as inpatient, outpatient, post-acute, and hospice spending at 1 year, did not differ between the two groups. These results suggest that MSU care may result in meaningful reductions in 1-year mortality among Medicare beneficiaries presenting with acute stroke symptoms, without increased health care utilization and costs.

Our study adds to the existing literature in several ways. First, it examines longer-term health, utilization, and spending outcomes for patients who received MSU care; most prior work focuses on outcomes during the index hospitalization and functional outcomes at 3 months. For example, a recent study found that mortality was somewhat lower at 90 days for MSU patients vs those transported by traditional EMS;^4^ other studies have found no significant difference in 7-day mortality.^7,17^ Second, this study is, to our knowledge, the first to link MSU registry data to claims data from the Medicare program, which covers more than 65 million Americans.^18^ Finally, the study expands the evidence base for MSU to a major metropolitan area—New York City, which is the largest and densest city in the US. Ambulance time and stroke care are known to differ across geographic areas, and it is important to understand how the clinical and financial impact of MSUs may vary accordingly.

Establishing an MSU involves significant upfront capital expenditures. Many hospitals have small operating margins and may hesitate to invest in the creation and maintenance of MSUs.^19^ Meanwhile, many payers do not currently reimburse for MSU services, and may resist doing so, without a clearer understanding of long-term health and financial outcomes.^20^ To our knowledge, no research has examined 1-year mortality differences for patients transported by MSUs vs traditional ambulances. With regard to spending, a recent study found that MSU care in Berlin was associated with higher costs relative to traditional care at 3 months;^21^ another analysis of patients mostly in the Houston area, which has not yet been peer-reviewed, reported similar costs across groups at 12 months.^22^ Both studies found that MSU care met commonly used cost-effectiveness thresholds.

Our study has limitations. First, this research examined outcomes for Medicare beneficiaries in a dense urban environment, and may not be generalizable to other populations, such as younger patients and patients in rural areas. Second, we were not able to directly examine the care that MSU and control patients received during hospitalization, such as acute blood control and antithrombotic and statin medications. Third, patients in the MSU registry were linked to Medicare claims through a series of steps and it is possible that some patients were misidentified; however, a sensitivity analysis with different matching criteria produced broadly similar results. Fourth, although MSU and control patients were well-balanced on various clinical and demographic characteristics, our results are subject to residual confounding. For instance, because of our reliance on Medicare claims data for our control group, we were unable to adjust for patients’ last known well time, which affects the odds of acute reperfusion therapies, and therefore could have impacted clinical outcomes. However, as the MSU generally only transports patients deemed eligible for acute reperfusion therapies, and more severe strokes tend to present earlier by EMS transport, we would expect such a selection bias to favor worse outcomes in the MSU group. Furthermore, because the study was observational, we cannot make causal claims about the impact of MSU care on patient outcomes.

Nonetheless, this study presents novel evidence on long-term outcomes for patients transported by MSU vs traditional ambulances in the largest city in the US. We find that MSU care was associated with a trend toward lower 1-year mortality for Medicare patients suspected to have acute stroke at presentation. Further research is needed to confirm these findings in other patient populations and regions of the country.

## Data Availability

Medicare claims can be accessed upon application to the Center for Medicare and Medicaid Services. EHR data can be accessed upon application to the Tripartite Request Assessment Committee (TRAC). The New York City mobile stroke registry is not available as it includes identifiable participant data and protected health information.

## Access to data

Dr Bond and Ms Pareek had full access to the data in the study and take responsibility for the integrity of the data and the accuracy of the data analysis.

